# NiaAge: a clinically interpretable measure of biological-age derived from long-term mortality-risk

**DOI:** 10.64898/2026.03.17.26348521

**Authors:** Jonah Kember, Emma Billington, Tanya ter Keurs, Monserrat Casado Sánchez, Mike Goss

## Abstract

Biological-age models quantify the physiological aging process by relating biomarker profiles (e.g., blood biochemistry, DNA methylation) to all-cause mortality risk. These models outperform chronological age in predicting disease and mortality, making them useful metrics for preventative health. However, in existing biological-age models, biomarker contributions do not align with the non-linear associations biomarkers exhibit with long-term mortality risk, nor do they account for normative trajectories that occur in healthy aging, limiting their utility in a clinical setting. To address these limitations, we developed a biological-age framework (NiaAge) where biomarker contributions are derived directly from non-linear associations with long-term mortality risk and aligned with normative trajectories observed in healthy aging. As a result, biomarker contributions to NiaAge are consistent with known biomarker risk profiles and normative reference ranges. We trained NiaAge in the 1999–2000 cohort of the US National Health and Nutrition Examination Survey (NHANES; *N*=2028) on 59 biomarkers spanning multiple physiological domains (e.g., hematology, metabolism, inflammation), then evaluated it in the 2001–2002 cohort (*N*=2346). NiaAge predicted long-term mortality, physical-health, and cognitive-health significantly better than chronological age. It also outperformed several DNA-methylation age clocks on mortality and physical/cognitive health-span metrics, while performing comparably to leading physiological age clocks. These results position NiaAge as a valuable tool for preventative health.

## Introduction

Aging is associated with a gradual decline in physiological function, yet the rate of this decline varies substantially across the population. To account for this variation, statistical models have been developed which estimate the aging process through direct physiological measurement, such as blood biochemistry (cholesterol, inflammatory markers, etc.) or DNA methylation (Kennedy et al., 2014; Levine et al., 2018; Fong et al., 2024). These models provide measures of biological-age, which are highly sensitive to the physiological aging process, surpassing chronological age in predicting the onset of disease, disability, and death (Li et al., 2025). This makes them highly desirable metrics in preventative health.

Contemporary models of biological-age, also known as age-clocks, are constructed by estimating the risk of all-cause mortality associated with a set of biomarker values, then identifying the chronological age for which that level of risk would be expected (Fong et al., 2024; Levine et al., 2018; Liu et al., 2019). In this framework, all-cause mortality is estimated as a linear combination of biomarkers through some form of statistical/machine learning model (e.g., PhenoAge, LinAge/LinAge2; Fong et al., 2024; Fong et al., 2025; Levine et al., 2018; Liu et al., 2019). To understand the specific biomarker values that are increasing and/or decreasing biological-age in these models, one can inspect how a biomarker profile maps onto a set of linear coefficients fitted during modelling.

When reporting biomarker results to patients, it is often beneficial to include normative reference ranges, clinical thresholds, and observed biomarker risk profiles (Van der Mee et al., 2024). Normative reference ranges indicate values that are typical for a patient’s age and sex, while clinical thresholds and biomarker risk profiles help contextualize whether deviations from the reference range are considered healthy or at-risk, helping guide treatment options. For a biological-age estimate to be clinically interpretable, the contribution of each individual biomarker should align with known risk profiles and normative (e.g., age- and sex-specific) reference ranges, to facilitate incorporation into the established reporting framework and shared decision-making process.

However, existing measures of biological-age suffer two limitations that make them difficult to reconcile with established clinical thresholds, demonstrated associations with risk, and normative reference-ranges. First, by assigning a single coefficient to each feature, existing age-clocks overlook the complex, often non-linear associations between biomarkers, all-cause mortality, and other relevant health outcomes (Nguyen et al., 2021; Clouston & Terrera, 2021). Instead, the contribution of a biomarker is simplified to ‘higher is better, lower is worse’, or vice-versa. Perhaps more problematic, since the strength and direction of biomarker contributions is heavily dependent on model hyper-parameters, rather than solely the biomarker-mortality association itself, it can vary widely between age-clocks, and even between versions of the same age-clock. For example, in PhenoAge, increased creatinine levels drive biological-aging upwards (*β=* .0095; per unit µmol/L); whereas in LinAge, they drive biological-age downwards (*β* = -.3536; per standardized unit relative to M±SD: 79.6 ± 13.2 *µ*mol/L). Studies modeling the shape of the creatinine–mortality association more precisely, however, suggest a non-linear relationship, with increased mortality-risk at very low (< 50 *µ*mol/L) and high creatinine levels (> 100 *µ*mol/L; Wang et al., 2024). Since non-linear biomarker–mortality associations are used when establishing clinical thresholds, this process results in an inherent discrepancy of biomarker contributions to biological-age with both clinical-thresholds and risk of all-cause mortality.

Second, existing biological-age models calculate biomarker-deviations relative to the sample mean/median, without considering how biomarker values naturally differ during typical aging. These models, therefore, may partially conflate increases in mortality-risk that arise during typical aging with increases that arise due to an underlying pathology. This also results in a discrepancy with the reference ranges that are typically provided in a clinical context, which in many cases take age into account. For example, an older adult might have a biomarker value that is exactly as expected for their chronological-age, yet this biomarker could still contribute substantially to their biological-age.

Finally, existing measures of biological-age typically rely on the Cox proportional hazards model (Fong et al., 2024; Fong et al., 2025; Levine et al., 2018; Liu et al., 2019), which explicitly assumes the biomarker-mortality association remains constant over the duration of the follow-up period (i.e., the 5-year hazard-ratio of a biomarker is equal to its 20-year hazard-ratio; Fox et al., 2002). However, this results in the underestimation of hazard-ratios for biomarkers in which abnormal levels disproportionately increase long-term relative to short-term mortality-risk. In biomarkers related to obesity, for instance, mortality-risk is significantly higher at long (>24 years) relative to short (< 12 years) follow-up durations, resulting in Cox models which underestimate the total cumulative impact of obesity on all-cause mortality over the lifespan (Haheime et al., 2007; Holme & Tonstad, 2013). To account for this, time-varying models, as well as cumulative statistics capable of summarizing these models, have been proposed to more accurately capture the biomarker–mortality association (Hernan et al., 2013).

To address these limitations, we developed a biological-age framework where the contribution of each biomarker to biological-age: (1) is derived directly from the non-linear association that a biomarker exhibits with all-cause mortality risk, (2) is aligned with biomarker-specific aging trajectories, and (3) accounts for long-term associations between biomarkers and mortality-risk. This measure of biological-age has several properties which increase its clinical interpretability. First, the contribution of each biomarker to biological-age is consistent with clinical thresholds and guidelines. Second, when a biomarker value is exactly as expected for an individual’s age and sex, its contribution to biological-age is exactly zero, which helps disentangle increases in mortality-risk that are due to normal-aging from those due to abnormal aging. As with other measures of biological-age, we maintain the property whereby any increase in biological-age has an equivalent effect on mortality-risk as aging that many years.

To evaluate performance, we evaluated our model against leading biological-age clocks derived from both DNA-methylation (PhenoAge, HannumAge, HorvathAge, GrimAge2Mort) and physiological measures (PhenoAge, LinAge/LinAge2, PCAge/PCAge2) using the same biomarkers and training/testing cohorts of the 1999–2001 US National Health and Nutrition Examination Survey (NHANES) as in prior work (Levine et al., 2018; Fong et al., 2024; Fong et al., 2025). We found that our measure of biological-age was a significantly stronger predictor of health and longevity than several DNA-methylation based age-clocks, and performed equally well compared to other leading physiological age-clocks.

## Results

### Development of a clinically interpretable biological-age framework

To construct a clinically interpretable measure of biological-age, we first modeled the univariate association between each biomarker and long-term mortality-risk. This was done through non-linear survival models, in line with contemporary work (Nguyen et al., 2021). To ensure sensitivity to long-term outcomes and to reduce bias from acute deaths, we used piecewise models with time-varying coefficients (Hernán et al., 2010). This approach allowed us to separately characterize short-term (e.g., 5-year) and long-term (e.g., 20-year) associations between biomarkers and mortality, shown in Figure 1A. On average, survival models showed a moderately non-linear time-varying effect (mean effective degrees of freedom in the smooth term: 4.7 ± 1.6; compared to a linear model with *df* = 1). To summarize these time-varying effects into a single long-term risk estimate, we computed the cumulative hazard across the full follow-up period (Hernán et al., 2010), shown in Figure 1B.

**Fig 1.**
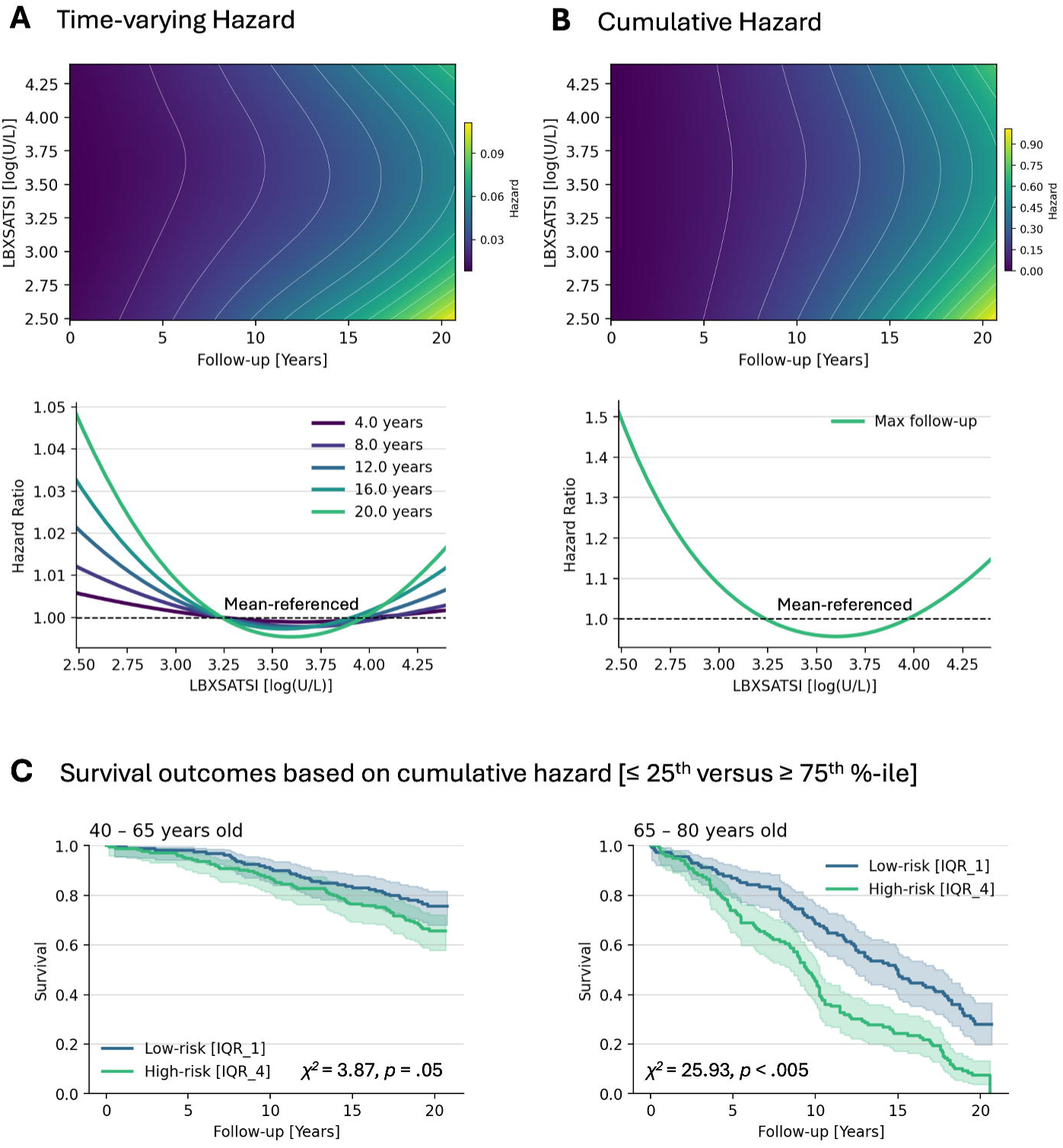
Modeling the association between biomarkers and long-term mortality risk. (A) Top: output of the time-varying survival model for Alanine Aminotransferase (ALT; NHANES-variable LBXSATSI); surface plot indicates 1-year hazard as a function of both follow-up time and LBXSATSI levels. Bottom: the hazard-ratio at specific follow-up durations (4, 8, 12, 16, 20 years), relative to the hazard expected at the mean LBXSATSI value within the sample. (B) Top: Cumulative hazard for the time-varying ALT survival model. Bottom: mean-referenced cumulative hazard-ratio curve. (C) Kaplan-Meier survival curves comparing subjects in the top and bottom quartiles based on cumulative hazard, calculated from the ALT model shown in panel B. On the left; 40–65 year olds. On the right; 65 - 80 year olds.

Next, we converted these hazard models into hazard-ratio models by referencing them to normative, age-expected values. These age-expected values were estimated using splines, which captured smooth, potentially non-linear associations between each biomarker and age. Together, this procedure yielded age- and biomarker-specific hazard-ratios, representing the relative change in long-term mortality risk associated with deviations from expected biomarker levels. Each biomarker hazard-ratio model was then rescaled such that a one-unit increase had an equivalent association with mortality-risk as aging one year; units referred to as Δ-age. This was done through the mortality-rate doubling time approximation (MRDT: = 7.5 years), as in the development of LinAge and LinAge2, where Δ-age is equal to ln(HR) / ln(2) * *MRDT* (Fong et al., 2024; Fong et al., 2025). An example of this referencing and conversion process for a single biomarker model can be seen in Figure 2.

**Fig 2.**
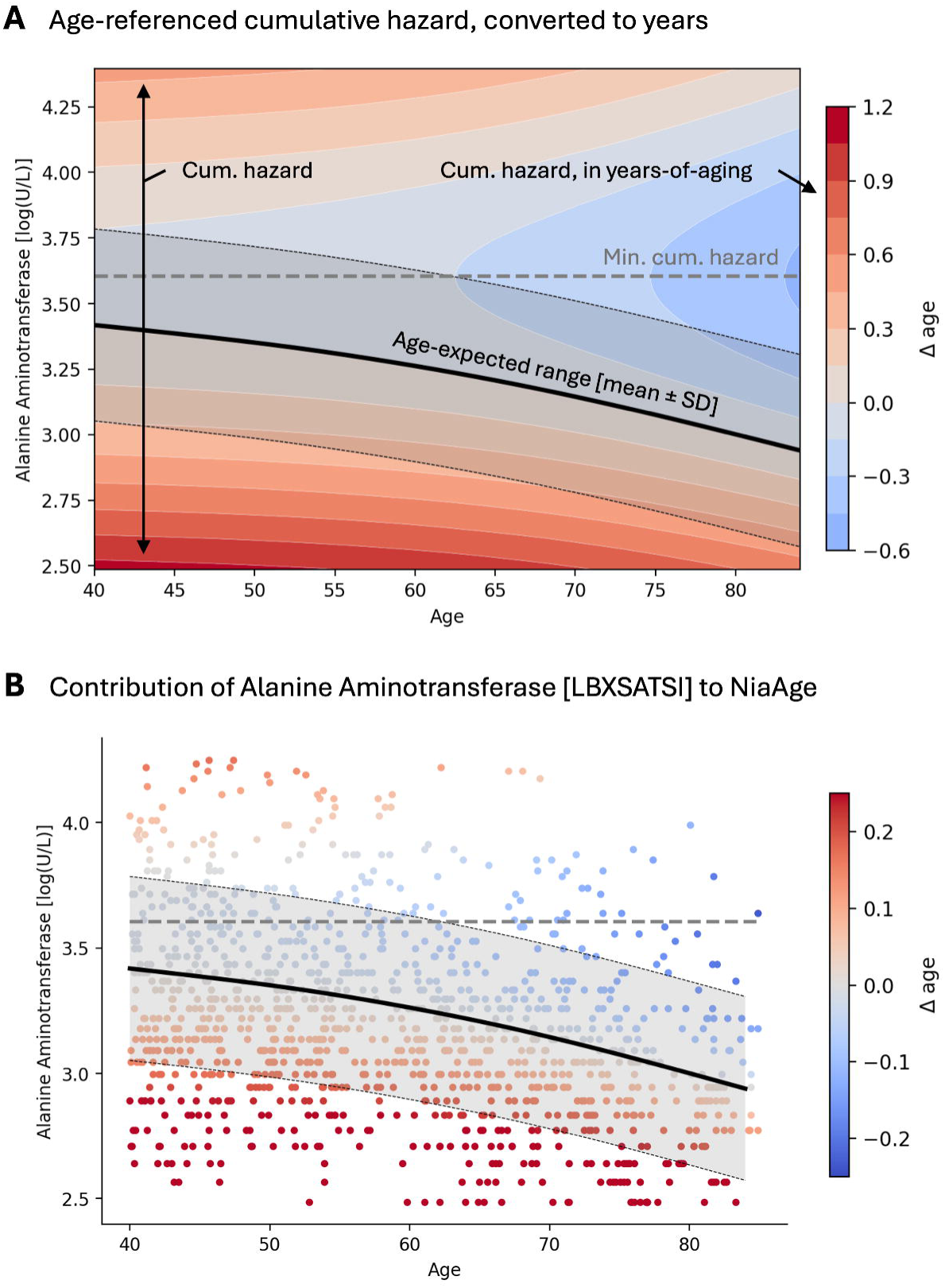
Example of single biomarker contribution to NiaAge. (A) An example biomarker model, where the contribution of a biomarker to NiaAge (Δ-age) varies as a function of both age and LBXSATSI levels. Color indicates the Δ-age for a specific LBXSATSI-age combination. The solid black line indicates the expected value of LBXSATSI given age, ± one standard deviation. (B) Scatterplot illustrating differences in LBXSATSI with age for each subject in the 1999–2000 NHANES cohort. Color indicates the delta-age, calculated from the model shown in panel A.

Finally, to summarize all of the risk-scores estimated from these biomarker models into a single composite score, we took a simple weighted sum. To preserve clinical interpretability, all weights were constrained to be non-negative, and were not optimized through multivariate modeling (which can produce sign flips in the presence of collinearity). Instead, we weighted each biomarker by its ability to predict mortality. Specifically, we evaluated the predictive performance of each biomarker model using the concordance index (*c*) and normalized these indices to derive biomarker weights (*w* = *c* / ∑*c*). We then applied a single scaling factor to these weights so that the summed, weighted score had an effect on long-term mortality that remained equivalent to one year of aging. This was done by fitting a Cox regression model on the composite risk- score, then using the MRDT approximation on the estimated hazard-ratio. A schematic illustrating this process is presented in Figure 3.

**Fig 3.**
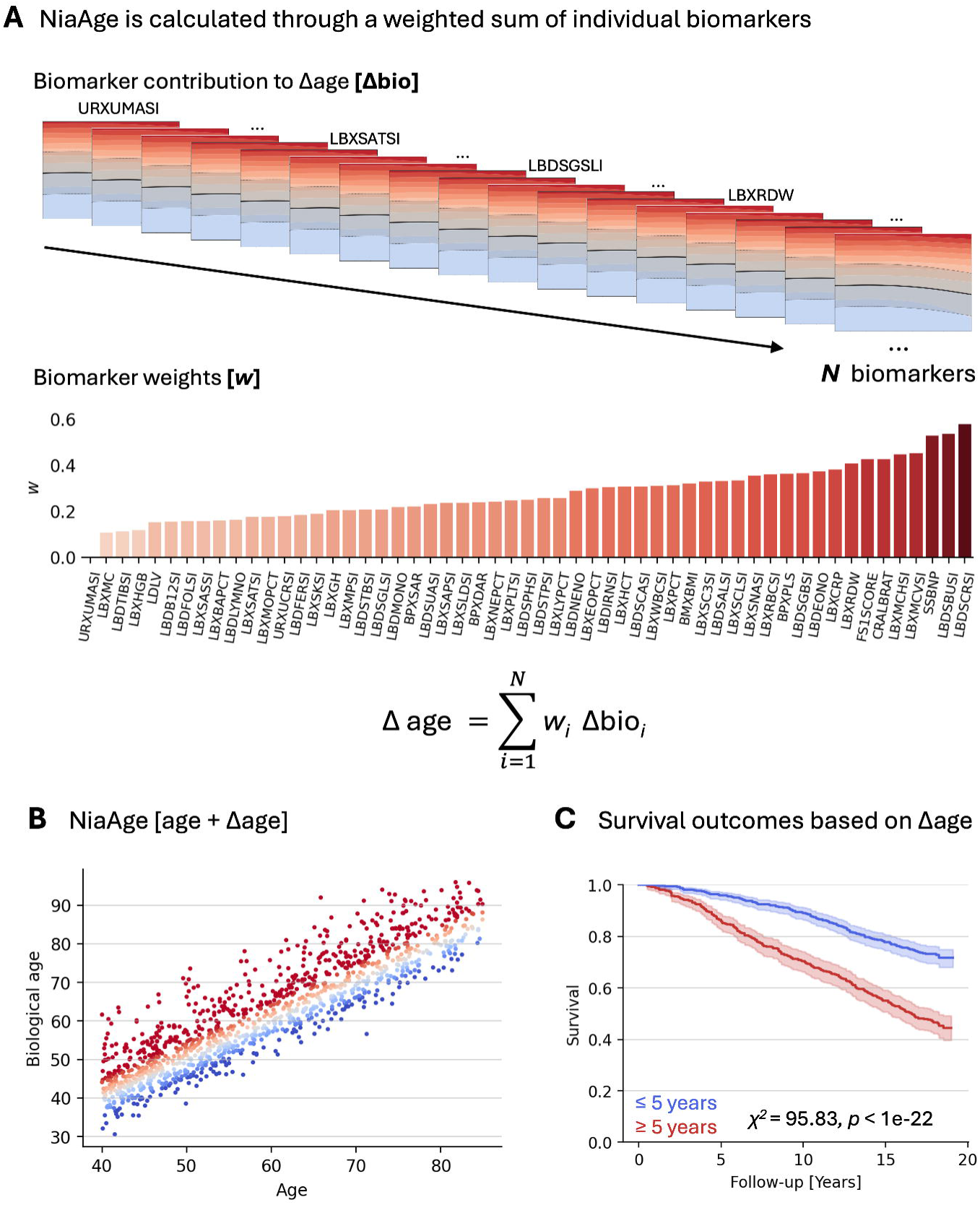
Calculating NiaAge. (A) Schematic illustrating how univariate biomarker models were aggregated to create NiaAge. Top: the univariate biomarker- and age-dependent risk models are all combined to create biological-age. Bottom: a non-negative weighting is applied to each based on, and these weights are re-scaled to maintain units that are equivalent in mortality-risk to aging. (B) Scatter-plot showing the final estimated NiaAge scores for all subjects (1999–2000). (C) Kaplan-Meier survival curves comparing subjects with a difference in chronological-age and NiaAge (Δ-age) greater than 5 years against subjects with a Δ-age less than 5 years.

All modeling was conducted separately in males and females. In males, the NHANES biomarkers with the highest weights, indicating the best mortality-prediction were: Creatinine (LBDSCRSI: *c* = .612; *p* < .005), Blood Urea Nitrogen (LBDSBUSI: *c* = .601; *p* < .005), NT-proBNP [surplus] (SSBNP, *c* = .599; *p* < .005), Mean cell volume (LBXMCVSI: *c* = .579; *p* < .005), and Mean cell hemoglobin (LBXMCHSI: *c* = .578; *p* < .005). In females, the biomarkers with the highest weights were: Serum Folate (LBDFOLSI: *c* = .607; *p* < .005), Bicarbonate (LBXSC3SI: *c* = .579; *p* < .005), Pulse (BPXPLS: *c* = .577; *p* < .005), NT-proBNP [surplus] (SSBNP: *c* = .575; *p* < .005), and Low-density Lipoprotein (LDLV: *c* = .563, *p* = .01). A full list is provided in the supplementary material.

### Interpreting the contribution of each biomarker to NiaAge

The primary advantage of our framework is that it allows the contribution of each biomarker to biological-age to be directly interpreted alongside normative references and clinical thresholds based on the known biomarker-risk profile. The contribution of each biomarker to biological-age is determined by the univariate association of that biomarker with long-term survival, as well as age/sex expected values. This results in two properties which lend clinical interpretability to the model. First, an individual with a biomarker value that is exactly as expected for their age and sex will always have a delta-age of exactly zero. Second, any deviation from that normative value will contribute to biological-age in a way that is directly aligned with the biomarker’s univariate association with long-term mortality risk.

These properties are illustrated in Figure 4. In panel A, *z*-score deviations from normative values highlight biomarkers for which this individual (a male in his 40’s) is higher or lower than expected for their age and sex. In panel B, biomarker contribution plots are equal to zero at exactly this age- and sex-expected value. For example, as shown, this individual, who has a biological age of 53, has an aspartate aminotransferase (AST) value of 3.89 log(U/L). This is 2.50 standard deviations higher than expected for his age (expected range: 3.23 ± 0.26 log(U/L)). Our biomarker model indicated that elevated AST increases this individual’s risk of long-term mortality. Accordingly, AST contributes +0.40 years to his biological age. The contribution of AST to biological age shown here aligns well with clinical reference ranges. Prior work indicates a similar “J-shaped” dose–response association between AST and all-cause mortality risk, with increased risk for AST levels above 3.31 log(U/L) and below 2.97 log(U/L), remarkably similar to what we observe here (Ling et al., 2024).

**Fig 4.**
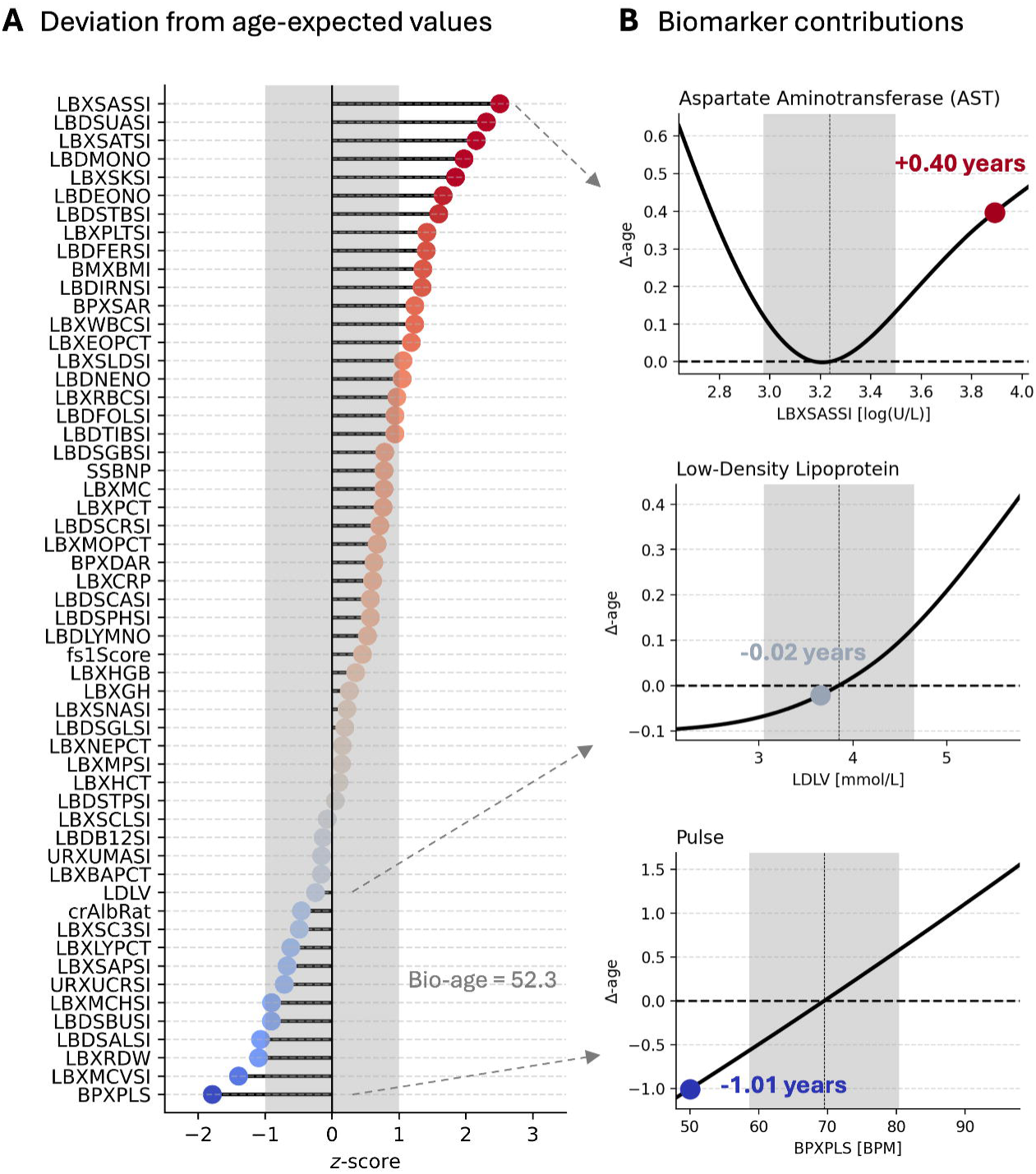
NiaAge is clinically interpretable: biomarker contributions align with long-term mortality-risk and age-norms. (A) Normative reference range for each biomarker, for a example subject (male in his 40’s). Dashed-line: expected value based on age/sex; shaded area: ± 1 standard deviation. Points: deviations from expected value for each biomarker. (B) Plots showing the contribution to biological-age as a smooth function of biomarker levels. Shaded-area: ± one standard deviation. Dashed-line: age-expected value, corresponding to a biological-age contribution of zero. Black line: contribution of biomarker to biological-age across a range of values.

**Fig 5.**
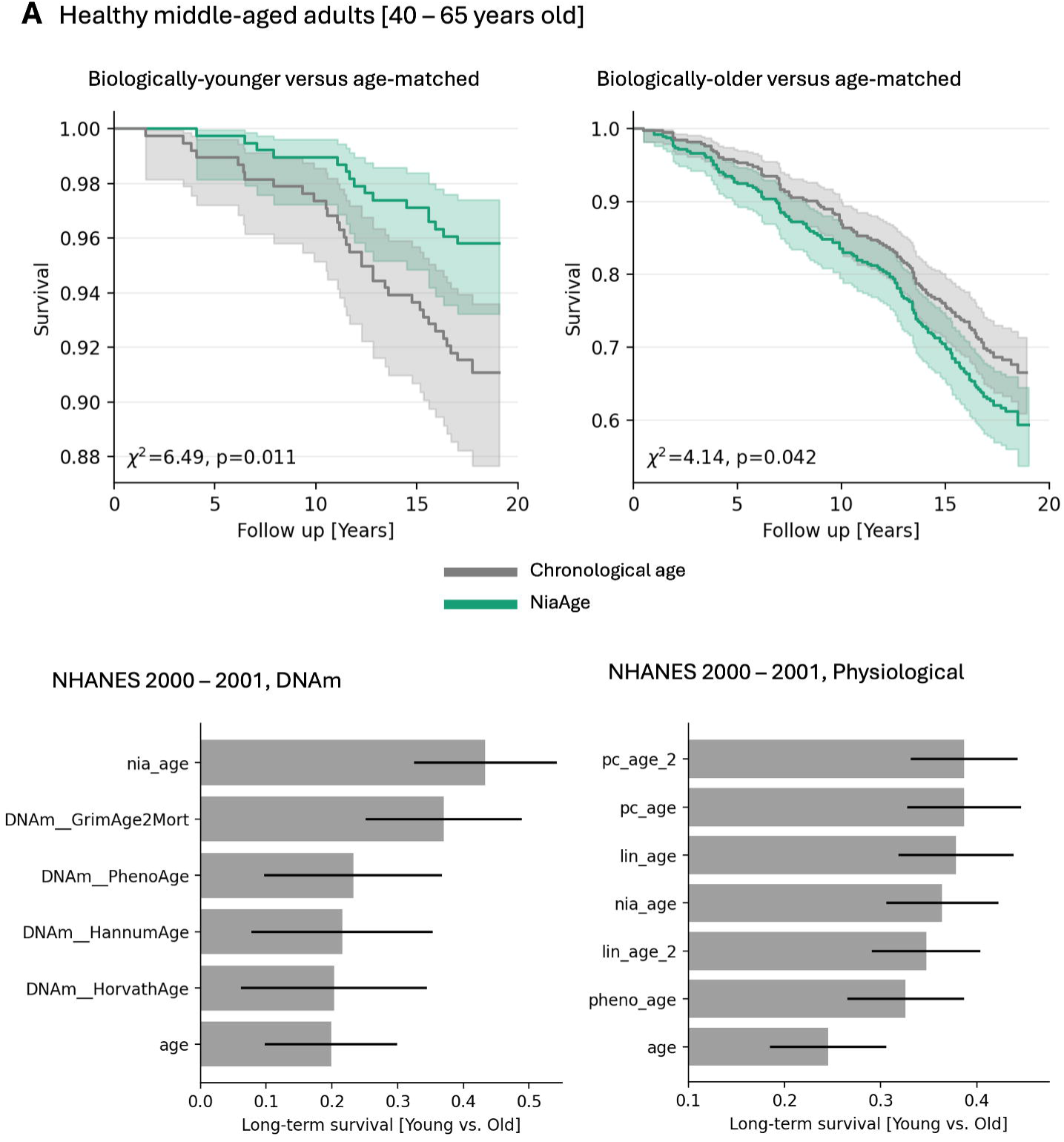
NiaAge is a strong predictor of long-term mortality. (A) Left: Kaplan-Meier curves comparing subjects in the bottom 25^th^ percentile of 40–65 year old subjects based on NiaAge (biologically-young) against those in the bottom 25^th^ percentile based on age (chronologically young). Right: same as right, comparing biologically and chronologically old subjects (> 75^th^ percentile). (B) Bar-plots indicate difference in survival probability at max follow-up (19 years) between older and younger subjects (<25^th^ vs. >75^th^ percentiles), as defined from each age-clock. Error bars indicate 95% confidence intervals.

**Fig 6.**
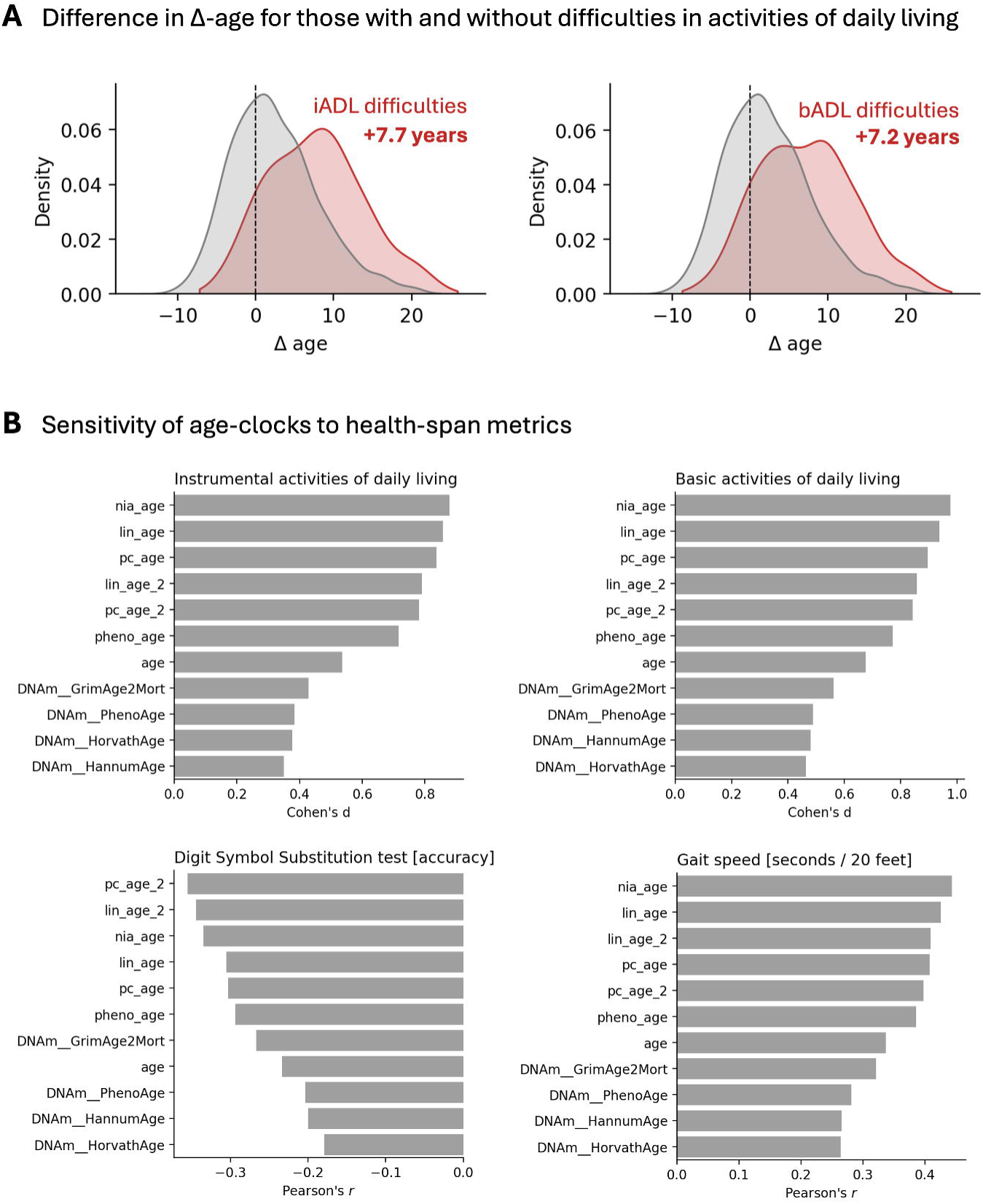
NiaAge is sensitive to health-span metrics. (A) Smoothed histograms (kernel density estimates) showing the distribution of subjects in the NHANES 2001–2002 cohort with and without difficulties in activities of daily living (red: with difficulties; gray: without difficulties). Left: difficulties with instrumental activities. Right: difficulties with basic activities. (B) Barplots comparing age-clocks in their sensitivity to health-span metrics. Top-left: Cohen’s d effect-size between those with and without iADL difficulties. Top-right: Cohen’s d effect-size between those with and without bADL difficulties. Bottom-left: Correlation between biological-age measures and accuracy on the Digit Symbol Substitution test. Bottom-right: Correlation between biological-age measures and gait-speed (time taken to cover 20 feet).

### Validating NiaAge against other leading markers

We evaluated NiaAge in its ability to predict long-term mortality, and benchmarked this performance against a set of widely used age-clocks. This included age-clocks calculated from physiological markers (LinAge, LinAge2, PCAge, PCAge2, PhenoAge) and those calculated from DNA-methylation (DNAm_PhenoAge, DNAm_HannumAge, DNAm_HorvathAge, DNAm_GrimAge2Mort; Levine et al., 2018; Horvath et al., 2013; Hannum et al., 2013; Lu et al., 2022). For fairness, we evaluated our model using the metrics selected by Fong et al. (2025) in their validation of LinAge2. All statistics are reported on the 2001–2002 NHANES test-set, which was not used in model fitting. Further details are provided in the methods.

First, we compared the Kaplan-Meier (KM) survival curves of subjects with high age-clock estimates (i.e., those identified as older; > 75^th^ percentile) between models through log-rank tests. In 65 to 74 year olds, we found that being biologically-older (high NiaAge) was a significantly stronger predictor of long-term survival when compared to being chronologically-older (*X*^2^ = 10.19, *p* < .005). Indeed, in biologically-older subjects, 20-year survival was 2.3 times lower compared to chronologically-older subjects (bio_age: 14.6%; age: 33.4%; difference: 18.8%). We also found that bio_age was a significantly stronger predictor of long-term survival when compared to DNAm HorvathAge (*X*^2^ = 4.72, *p* = .03). We found no significant difference between bio-age and any other age-clocks (all *p’s >* .05). Similarly, in 40 to 65 year olds, we found that being biologically-older (high NiaAge) was a significantly stronger predictor of long-term survival when compared to being chronologically-older (*X*^2^ = 6.34, *p* = .01). Indeed, in biologically-older individuals, 20-year survival was 7.2% lower compared to chronologically-older individuals (bio_age: 59.3%, age: 66.5%). We also found that NiaAge was a significantly stronger predictor of long-term survival when compared to: DNAm HannumAge (*X*^2^ = 5.18, *p* = .02) and DNAm HorvathAge (*X*^2^ = 4.72, *p* = .03). We found no significant difference between bio-age and any other age-clocks (all *p’s > .05*).

We also compared the KM survival curves of subjects with low age-clock estimates (i.e., those identified as younger) between models. In 65 to 74 year olds, we found that being biologically-younger (low NiaAge) was a significantly stronger predictor of long-term survival when compared to being chronological-younger (*X*^2^ = 7.90, *p* < .005). Indeed, in biologically-younger individuals, 20-year survival was 15.6% higher compared to chronologically-younger individuals (bio_age: 73.3%; age: 57.7%). We found no significant difference between NiaAge and any other age-clocks (all *p’s* > .05). In 40 to 65 year olds, we found that being biologically-younger was a significantly stronger predictor of long-term survival when compared to being chronological-younger (*X*^2^ = 4.14, *p* = .04). Indeed, in biologically-younger subjects, 20-year survival was 4.6% higher compared to chronologically-younger subjects (95.8% versus 91.2%). We also found that bio_age was a significantly better predictor of long-term survival compared to DNAm PhenoAge (X^2^ = 4.38, *p* = .04). We found no significant difference between NiaAgeand any other age-clocks (all *p’s* > .05).

### Evaluating the sensitivity of NiaAge to health metrics important for longevity

Next, we evaluated the sensitivity of NiaAge to measures of physical and cognitive health, and compared this sensitivity to other age-clocks. First, we found that individuals with difficulties in the ability to perform Basic Activities of Daily Living (bADL) had a biological-age that was 13.17 years older than individuals without any bADL difficulties, a large effect-size significant difference (*t*(270) = 14.65, *p* < 1e-36, Cohen’s *d* = .97). This difference was significantly larger than that of chronological-age (Cohen’s *d* = .67; bio-age vs. age: *p* = .00013) and all DNA methylation age-clocks (max Cohen’s *d*: .56; max bio-age vs. DNAm-age *p-*value = .0012). It was not significantly different when compared to that of the other physiological age-clocks (min bio-age versus age-clocks: *p* = .22).

Similar effects were observed when contrasting subjects with and without difficulties in the ability to perform Instrumental Activities of Daily Living (iADL). Indeed, subjects with iADL difficulties had a biological-age that was 11.98 years older than subjects without any iADL difficulties, a significant difference (*t*(221) = 11.5, *p* = 1e-24, Cohen’s *d* = .88). This difference was significantly larger than that of chronological-age (Cohen’s *d* = .54; bio-age vs. age: *p* = .0002) and all DNA methylation age-clocks (max-*p* = .0015). It was not significantly different when compared to that of the other physiological age-clocks (min-*p* = .35).

Next, we examined scores on the Digital Symbol Substitution Test (DSST). This test captures response speed, sustained attention, visual spatial skills, associative learning, and memory; and is thought to be sensitive to dementia (Jaeger et al., 2018). We found a negative correlation between biological-age and DSST scores (*r* = -.33, *p* < 1e-23; correlation with delta-aga: *r* = -.24, *p* < 1e-15), indicating better performance in those identified as biologically-younger. This was significantly stronger than the correlation between DSST scores and chronological age (*r* = -.23; bio_age vs. age: *p* = .01). This was also significantly stronger than the correlation between DSST scores and DNAm_HorvathAge (difference-*p* = .036), and trended towards a significantly stronger correlation compared to DNAm_HannumAge (*p* = .08). This was not significantly different from any of the other age-clocks (min-*p* = .11).

Finally, we examined gait-speed, a measure of physical function that slows with aging. We found a strong, significant correlation between biological-age and gait-speed (*r* = .44, *p* < 1e-75; correlation with delta-aga: *r* = .32, *p* < 1e-38), indicating faster gait-speed in those identified as biologically-younger. This was significantly stronger than the correlation between gait-speed scores and chronological age (*r* = .33; bio_age vs. age: *p* = .0005). This was also significantly stronger than the correlation between gait-speed and all DNA-methylation age-clocks (difference-*p* = .036), and trended towards a significantly stronger correlation for pheno_age (bio_age vs. pheno_age: *p* = .064). This was not significantly different from any of the other age-clocks (min-*p* = .11).

## Discussion

Measures of biological age are increasingly used in preventative healthcare to improve upon chronological age in capturing the physiological aging process. These age-clocks are highly sensitive to declines in physiological function, as reflected in their ability to predict mortality and their sensitivity to health and longevity metrics. However, many age-clocks lack clinical interpretability, limiting the extent to which they can be incorporated alongside clinical thresholds and normative reference ranges. To address this, we constructed a framework for measuring biological age that: (1) is directly tied to the univariate dose-response association that biomarkers show with all-cause mortality across the lifespan, (2) accounts for age- and sex-expected reference ranges, and (3) accounts for long-term associations between biomarkers and mortality-risk.

We then constructed a measure of biological age by applying this framework to the NHANES dataset, and evaluated this biological-age clock against existing age-clocks derived from DNA methylation and physiological markers. We found our measure of biological age to be a strong predictor of long-term health outcomes, with high sensitivity to physical and cognitive health metrics related to longevity. Our measure of biological age outperformed chronological age across all examined outcomes, and performed significantly better than several DNA methylation-based age-clocks. We also found no significant difference in the ability to predict long-term mortality between our measure of biological age and a set of commonly used physiological age-clocks. Numerically, our measure of biological age was also the single most sensitive age-clock across all physical health metrics examined (iADL, bADL, gait speed).

To highlight the clinical interpretability of this framework, we provided the biomarker contributions to biological-age alongside normative reference ranges for a male in his 40’s. For each biomarker, there is a logical consistency between: deviations from normative reference values based on age and sex (e.g., +1.5 standard deviations), biomarker contributions to biological-age (e.g., + 0.5 years), and biomarker–mortality associations (e.g., dose-response curves with age/sex expected values equal to zero). This allows patients to readily identify the specific biomarkers for which they exhibit abnormal levels (i.e., different from their age-expected value), and understand whether these abnormalities may be of concern. Meta-analytical evidence suggests that indicating how a biomarker result compares to what is expected for age increases patient understanding of lab results (Van der Mee et al., 2024).

An important advantage of our framework is its emphasis on modeling the univariate dose-response association between individual biomarkers and long-term mortality risk. New biomarkers relevant to healthspan are regularly discovered, and existing associations are regularly refined (Clouston & Tererra, 2021). By emphasizing univariate biomarker–healthspan relationships, our framework readily supports the incorporation of new biomarkers without disrupting existing biomarker–mortality associations. Moreover, by modeling the biomarker-risk profile across the entire range of the biomarker, as opposed to using a coarse division of the biomarker into normal/abnormal ranges (e.g., percentile-based cutoffs; Liu et al., 2020), our framework allows for the interpretation of biomarker-contributions to biological-age even when values fall within the expected range.

It should be noted that while models such as PhenoAge, LinAge, or LinAge2 do not have univariate alignment with biomarker-mortality associations, limiting their clinical interpretability, this does not represent an inherent limitation of their ability to predict mortality. As shown in our model comparisons, the predictive performance of these models is quite high. They were simply not designed with direct clinical interpretability as a primary objective. In fact, to address this issue, Fong et al. (2025) provide extensive detail on the principal components underlying PCAge, along with highly detailed instructions on their interpretation. However, interpreting these component scores safely requires increased statistical expertise, and the framework has received limited external validation or mainstream adoption that would facilitate broader clinical use.

## Limitations

Our biological-age framework is not without limitations. First, our framework is limited in its ability to capture multivariate effects, where combinations of biomarker values contribute to mortality risk beyond the sum of their individual effects. Instead, when specific interactions are known to impact mortality-risk, they must be included as separate biomarkers (e.g., Albumin:Creatinine ratio, ApoB/ApoA1 ratio). Second, as with all measures of biological-age, a crucial limitation is the correlational nature of the underlying survival models: the associations we report between biomarkers and outcomes reflect statistical relationships, rather than causal mechanisms. These associations should therefore not be interpreted as if modifying a given biomarker will directly alter mortality risk. Third, the data-set used here, and in the development of other biological-age clocks, relies on sampling physiological measurements at single time points. While evidence indicates many of these single time-point measures remain predictive over long follow-up durations (Haheim et al., 2007), it is in many instances the variability of the measures, as well as the cumulative exposure to abnormal levels, that help accurately predict mortality-risk (e.g., estimated glomerular filtration rate; Al-Aly et al., 2012). Future biological-age clocks will benefit from data-sets which better incorporate variability in physiological measurements across time into their risk estimates. Finally, spline-based models can be less reliable at the boundaries of the observed data, where sample size is limited, leading to lower stability in the estimated parameters.

## Conclusions

Here we developed and validated NiaAge, a biological-age framework based on the non-linear associations biomarkers exhibit with long-term mortality-risk, referenced to typical aging trajectories. The purpose of this framework is to facilitate patient understanding of individual biomarker results in clinical practice. NiaAge does so by ensuring that: (1) biomarker values which are exactly as expected for one’s age and sex contribute exactly zero years to the final biological-age estimate, and (2) any deviation from the age-expected biomarker value contributes to biological-age in a way that is aligned with the biomarker–mortality risk profile.

## Methods

### Data

For modeling, we used data from the US National Health and Nutrition Examination Survey, a nationally representative study that collects health, demographic, and physiological measurements from the civilian population. We used the 1999–2000 sample for training (*N*=2082; 1047 male, 1035 female) and the 2000–2001 sample for testing (*N*=2346; 1202 male, 1144 female). To ensure differences in model performance relative to existing biological-age clocks were not driven by differences in low-level preprocessing steps, we used the data preprocessed by Fong et al. (2025; available in their supplementary material: mergedDataNHANES9902.csv). This data was filtered to subjects aged 40–84 years old, and had a subset of biomarkers log-transformed (NHANES variables: BMXBMI, LBDFERSI, LBDFOLSI, LBDB12SI, LBDSCRSI, LBXCRP, LBXSAPSI, LBXSASSI, LBXSATSI, LBXSLDSI, LBXWBCSI, SSBNP, crAlbRat).

### Model fitting

Survival modeling was done independently on each biomarker, for males and females separately. For each biomarker, we first removed outlier values beyond the 2.5^th^ and 97.5^th^ percentiles prior to fitting. Prior work indicates this helps mitigate the tendency for splines to overfit data at the tails of the distribution (Nguyen et al., 2021). Next, we prepared data for piecewise-exponential modeling by converting from wide-format to long-format using 1-year intervals. We then fitted non-linear, non-linearily time-varying models using the standard pipeline in the R-package *pammtools* (Bender et al., 2018; example: https://adibender.github.io/pammtools/articles/tveffects.html). This was implemented through a tensor product term between the biomarker and piecewise follow-up time (model parameters: family=poisson; gamma=1; method=REML). All models included age as a covariate.

To derive age-expected values, we predicted each biomarker from age using Generalized Additive Models (GAMs, LinearGam function from the pygam package; Serven et al., 2018). These GAMs used a Normal error distribution and identity link, with all parameters at default except lambda, which was increased to 1e3 to prevent overfitting. Visual inspection confirmed that this produced smooth age–biomarker associations with relatively little bias. Age-expected values were then used to re-reference the cumulative hazard curves, ensuring that all measures of cumulative hazard are expressed relative to the age-expected hazard.

Once we had: (1) modeled the non-linear, univariate association of each biomarker with cumulative hazard, (2) referenced these hazard estimates relative to age-expected values, and (3) converted these hazard-ratios to units of Δ-age, we combined these scores into a single composite score. To ensure that the direction and relative contribution of each biomarker to this composite score was consistent with its univariate association, we used a simple weighted combination rather than fitting a multivariate model (Song et al., 2017). To derive biomarker-weights, we quantified the ability of each biomarker-model to predict mortality using the concordance-index, then normalized these concordance indices across biomarkers. Finally, we rescaled the composite risk score to express changes in terms of equivalent years-of-aging. Specifically, we fitted a Cox regression model on the composite risk-score, calculated the number of hazard doublings via ln(HR)/ln(2), then converted this into years assuming a mortality-rate doubling time of 7.5 years.

### Model evaluation

We evaluated our biological-age measure in its ability to predict long-term mortality, and its sensitivity to healthspan. To ensure we conducted a fair comparison against existing age-clocks: our training and testing splits were defined on the same NHANES subjects, our model fitting was done using the same set of *N*=59 physiological biomarkers (a full list is provided in the Supplementary material), and we used the exact model evaluation criteria described by Fong et al. (2025). When predicting our measure of biological-age in the test-set cohort, biomarker values were clipped to the 2.5^th^ and 97.5^th^ percentiles of the training data.

To evaluate long-term mortality, we stratified each age-clock into its highest and lowest quartiles (<25^th^ percentile; >75^th^ percentile), then compared the Kaplan-Meier curves of those quartiles between age-clocks using log-rank tests. Each test was done separately for specific age-bins: 40–65 year olds and 65–74 year olds and evaluated on both training (NHANES 1999–2000) and testing data (NHANES 2001–2002), as in Fong et al. (2024).

To evaluate sensitivity to healthspan, we related age-clocks to measures of physical and cognitive function. For physical function, we identified subjects who have difficulties with basic activities of daily living (bADL), defined as having responded “some difficulty” or higher on any of the Physical Functioning questions: [PFQ060B, PFQ060C, PFQ060H, PFQ060I, PFQ060J, PFQ060K, PFQ060L] (e.g., PFQ060C: “By yourself and without using any special equipment, how much difficulty do you have walking up 10 steps without resting?”). We also measured Gait speed, defined as the time taken to walk 20 feet. To measure cognitive function, we used scores on the Digital Symbol Substitution Test (DSST, NHANES variable CFDRIGHT), a validated measure of cognitive processing speed on the Wechsler Adult Intelligence (IQ) scale (Jaeger et al., 2018).

For all tests, we compared our measure of biological-age to: chronological-age, modern physiology-based age-clocks (LinAge, LinAge2, PCAge, PCAge2, PhenoAge), as well as DNA-methylation age-clocks (DNAm_PhenoAge, DNAm_HannumAge, DNAm_HorvathAge, DNAm_GrimAge2Mort; Levine et al., 2018; Horvath et al., 2013; Hannum et al., 2013; Lu et al., 2022). Each of these physiological measures of biological-age was developed using the exact same train/test set subjects and biomarkers as our measure, making these measures directly comparable. Only a subset of NHANES subjects have DNA-methylation data (*N*=1019 in the 1999–2000 cohort; *N*=1065 in the 2000–2001 NHANES cohort). These subjects were therefore matched in any statistical tests comparing biological-age against DNAm-clocks.

## Data availability

NHANES data is openly available through the Center for Disease Control and Prevention (CDC) website, https://wwwn.cdc.gov/nchs/nhanes/. Cleaned data used in analyses is provided in the supplement.

## Code availability

All code required to fit our measure of biological-age is available at [https://github.com/JonahKember/biological_age]. All code necessary to re-create analyses is available at: [https://github.com/JonahKember/biological_age_analyses].

