## Supplementary material for "NiaAge: a clinically interpretable measure of biological-age derived from long-term mortality-risk"

**Table 1.** NiaAge male biomarker weights

| **biomarker** | **biomarker_label** | **weight__male** |
|---|---|---|
| LBDSCRSI | Creatinine | 1 |
| LBDSBUSI | Blood Urea Nitrogen | 0.926844553 |
| SSBNP | NT-proBNP (surplus) | 0.913494838 |
| LBXMCVSI | Mean cell volume | 0.779098915 |
| LBXMCHSI | Mean cell hemoglobin | 0.76977308 |
| crAlbRat | Log Urine Albumin-to-Creatinine Ratio | 0.737375903 |
| fs1Score | Co-morbidity index. | 0.734646591 |
| LBXRDW | Red cell distribution width | 0.702521521 |
| LBXCRP | CRP | 0.660402718 |
| LBDEONO | Eosinophils number | 0.644867905 |
| LBDSGBSI | Globulin | 0.629704146 |
| BPXPLS | Pulse | 0.625672021 |
| LBXRBCSI | Red blood cell count | 0.622967446 |
| LBXSNASI | Sodium | 0.611604934 |
| LBXSCLSI | Chloride | 0.575991952 |
| LBDSALSI | Albumin | 0.570219994 |
| LBXSC3SI | Bicarbonate | 0.568298757 |
| BMXBMI | Body Mass Index | 0.552005343 |
| LBXPCT | Transferrin Saturation | 0.540444936 |
| LBXWBCSI | WBC count | 0.535258419 |
| LBDSCASI | Calcium total | 0.529247337 |
| LBXHCT | Hematocrit | 0.529008213 |
| LBDIRNSI | Iron | 0.526781886 |
| LBXEOPCT | Eosinophils | 0.518066229 |
| LBDNENO | Segmented neutrophils number | 0.500494739 |
| LBXLYPCT | Lymphocyte | 0.445875524 |
| LBDSTPSI | Protein total | 0.442634981 |

| | | |
|---|---|---|
| LBDSPHSI | Phosphorus | 0.430142815 |
| LBXPLTSI | Platelet count | 0.427990699 |
| LBXNEPCT | Segmented neutrophils | 0.416636433 |
| BPXDAR | Diastolic blood pressure | 0.412051849 |
| LBXSLDSI | Lactate Dehydrogenase (LDH) | 0.407994987 |
| LBXSAPSI | Alkaline Phosphatase (ALP) | 0.406321119 |
| LBDSUASI | Uric acid | 0.400780039 |
| BPXSAR | Systolic blood pressure | 0.380124674 |
| LBDMONO | Monocyte number | 0.376389393 |
| LBDSGLSI | Glucose | 0.359931066 |
| LBDSTBSI | Bilirubin total | 0.357976846 |
| LBXMPSI | Mean platelet volume | 0.352237871 |
| LBXGH | Glycohemoglobin | 0.351660675 |
| LBXSKSI | Potassium | 0.327138098 |
| LBDFERSI | Ferritin | 0.318290511 |
| URXUCRSI | Creatinine urine | 0.307958706 |
| LBXMOPCT | Monocyte | 0.303819387 |
| LBXSATSI | Alanine Aminotransferase (ALT) | 0.302582539 |
| LBDLYMNO | Lymphocyte number | 0.280434711 |
| LBXBAPCT | Basophils | 0.275215212 |
| LBXSASSI | Aspartate Aminotransferase (AST) | 0.272246776 |
| LBDFOLSI | Folate serum | 0.272139582 |
| LBDB12SI | Vitamin B12 serum | 0.268626934 |
| LDLV | Low-Density Lipoprotein | 0.264767967 |
| LBXHGB | Hemoglobin | 0.205828029 |
| LBDTIBSI | Total iron binding capacity | 0.194325341 |
| LBXMC | Mean Cell Hemoglobin Concentration | 0.185024242 |
| URXUMASI | Albumin urine | 0 |

**Table 2.** NiaAge female biomarker weights

| **biomarker** | **biomarker_label** | **weight__female** |
|---|---|---|
| LBDFOLSI | Folate serum | 1 |
| fs1Score | Co-morbidity index. | 0.951092012 |
| LBXSC3SI | Bicarbonate | 0.781103197 |
| BPXPLS | Pulse | 0.766315418 |
| SSBNP | NT-proBNP (surplus) | 0.752069229 |
| crAlbRat | Log Urine Albumin-to-Creatinine Ratio | 0.703867664 |
| LDLV | Low-Density Lipoprotein | 0.656972979 |
| LBXRDW | Red cell distribution width | 0.634579384 |
| LBXPLTSI | Platelet count | 0.61847295 |
| URXUCRSI | Creatinine urine | 0.581056102 |
| LBDSTBSI | Bilirubin total | 0.55886266 |
| BPXSAR | Systolic blood pressure | 0.542650262 |
| LBXWBCSI | WBC count | 0.510025313 |
| LBXSASSI | Aspartate Aminotransferase (AST) | 0.505245187 |
| LBXCRP | CRP | 0.488962148 |
| LBDNENO | Segmented neutrophils number | 0.444798964 |
| LBDTIBSI | Total iron binding capacity | 0.426267145 |
| LBXSKSI | Potassium | 0.404886089 |
| LBDIRNSI | Iron | 0.404650615 |
| LBDSUASI | Uric acid | 0.388850297 |
| LBDMONO | Monocyte number | 0.380844175 |
| LBDSBUSI | Blood Urea Nitrogen | 0.380314358 |
| BPXDAR | Diastolic blood pressure | 0.376452581 |
| LBDEONO | Eosinophils number | 0.376052275 |
| LBDSCRSI | Creatinine | 0.374486372 |
| LBXLYPCT | Lymphocyte | 0.356225349 |
| LBXSAPSI | Alkaline Phosphatase (ALP) | 0.35019721 |

| | | |
|---|---|---|
| LBXNEPCT | Segmented neutrophils | 0.345876258 |
| LBDLYMNO | Lymphocyte number | 0.322340613 |
| LBXSLDSI | Lactate Dehydrogenase (LDH) | 0.319562018 |
| LBDSGLSI | Glucose | 0.309142285 |
| LBDSPHSI | Phosphorus | 0.297768882 |
| LBXSCLSI | Chloride | 0.296885854 |
| LBDFERSI | Ferritin | 0.296826985 |
| LBXSNASI | Sodium | 0.293082946 |
| LBXRBCSI | Red blood cell count | 0.291269795 |
| LBXHGB | Hemoglobin | 0.286313063 |
| LBXHCT | Hematocrit | 0.282757403 |
| LBDSGBSI | Globulin | 0.277706481 |
| LBXMC | Mean Cell Hemoglobin Concentration | 0.263954789 |
| LBDB12SI | Vitamin B12 serum | 0.263142403 |
| LBXMPSI | Mean platelet volume | 0.261623595 |
| LBXEOPCT | Eosinophils | 0.260834756 |
| LBDSALSI | Albumin | 0.258173898 |
| LBXMCVSI | Mean cell volume | 0.257090716 |
| LBXMOPCT | Monocyte | 0.254900806 |
| LBDSTPSI | Protein total | 0.228374639 |
| LBDSCASI | Calcium total | 0.214846647 |
| BMXBMI | Body Mass Index | 0.193724613 |
| LBXMCHSI | Mean cell hemoglobin | 0.180184847 |
| LBXBAPCT | Basophils | 0.112085713 |
| LBXSATSI | Alanine Aminotransferase (ALT) | 0.111143816 |
| LBXPCT | Transferrin Saturation | 0.083275446 |
| URXUMASI | Albumin urine | 0.06941779 |
| LBXGH | Glycohemoglobin | 0 |

**Figure 1.** NiaAge is highly correlated with existing biological age clocks

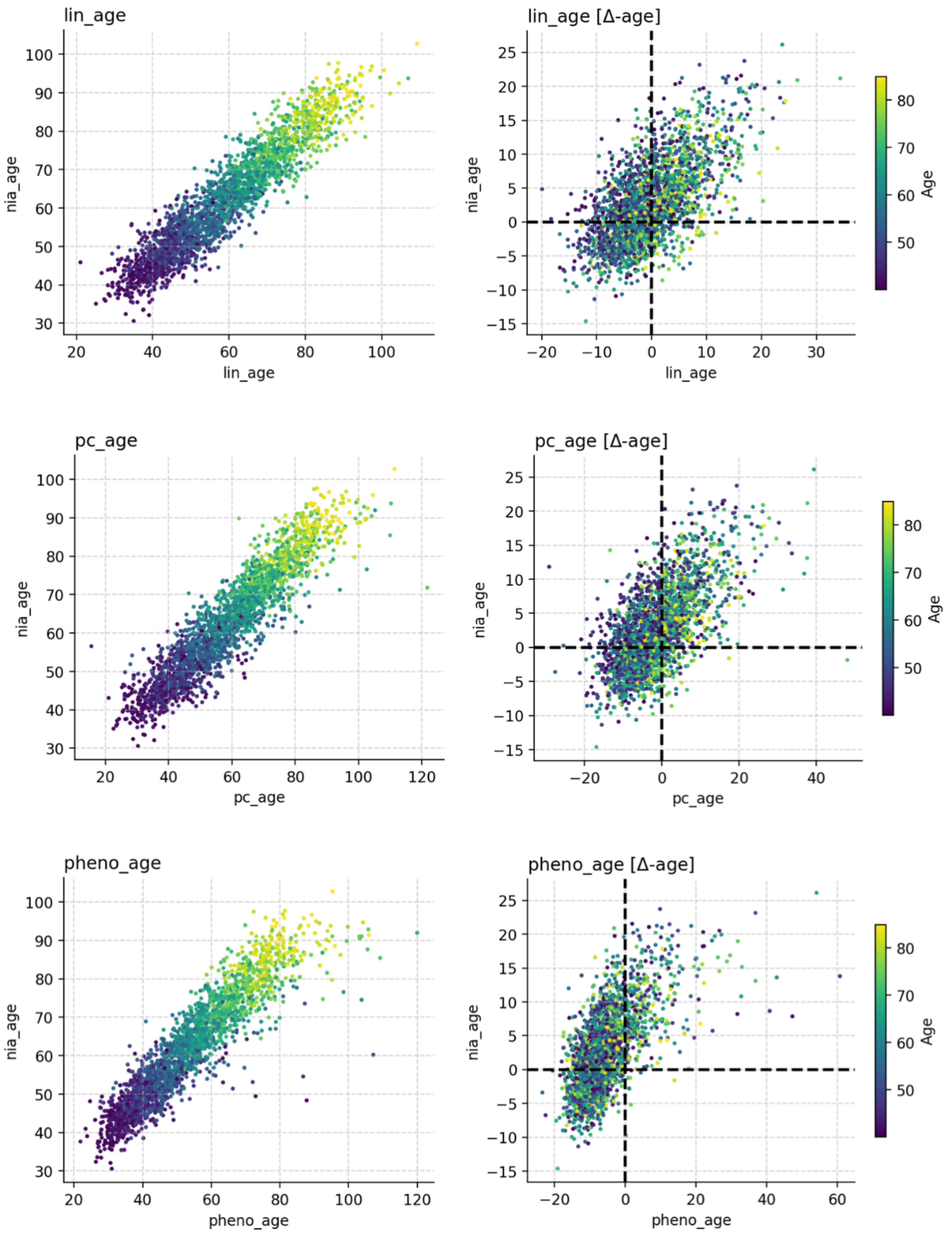

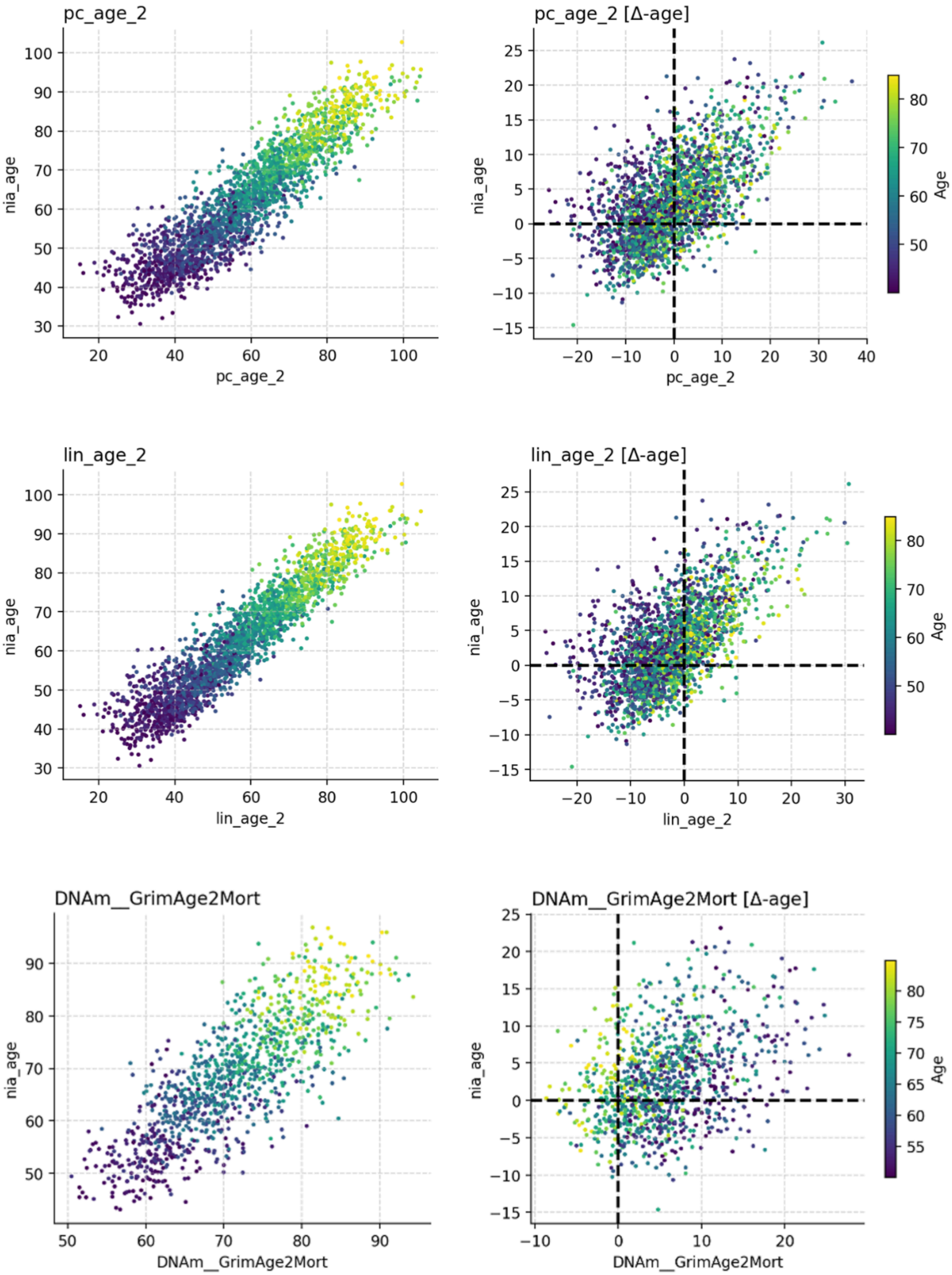
pc_age_2
nia_age
pc_age_2
pc_age_2 [Δ-age]
nia_age
pc_age_2
Age
lin_age_2
nia_age
lin_age_2
lin_age_2 [Δ-age]
nia_age
lin_age_2
Age
DNAm__GrimAge2Mort
nia_age
DNAm__GrimAge2Mort
DNAm__GrimAge2Mort [Δ-age]
nia_age
DNAm__GrimAge2Mort
Age

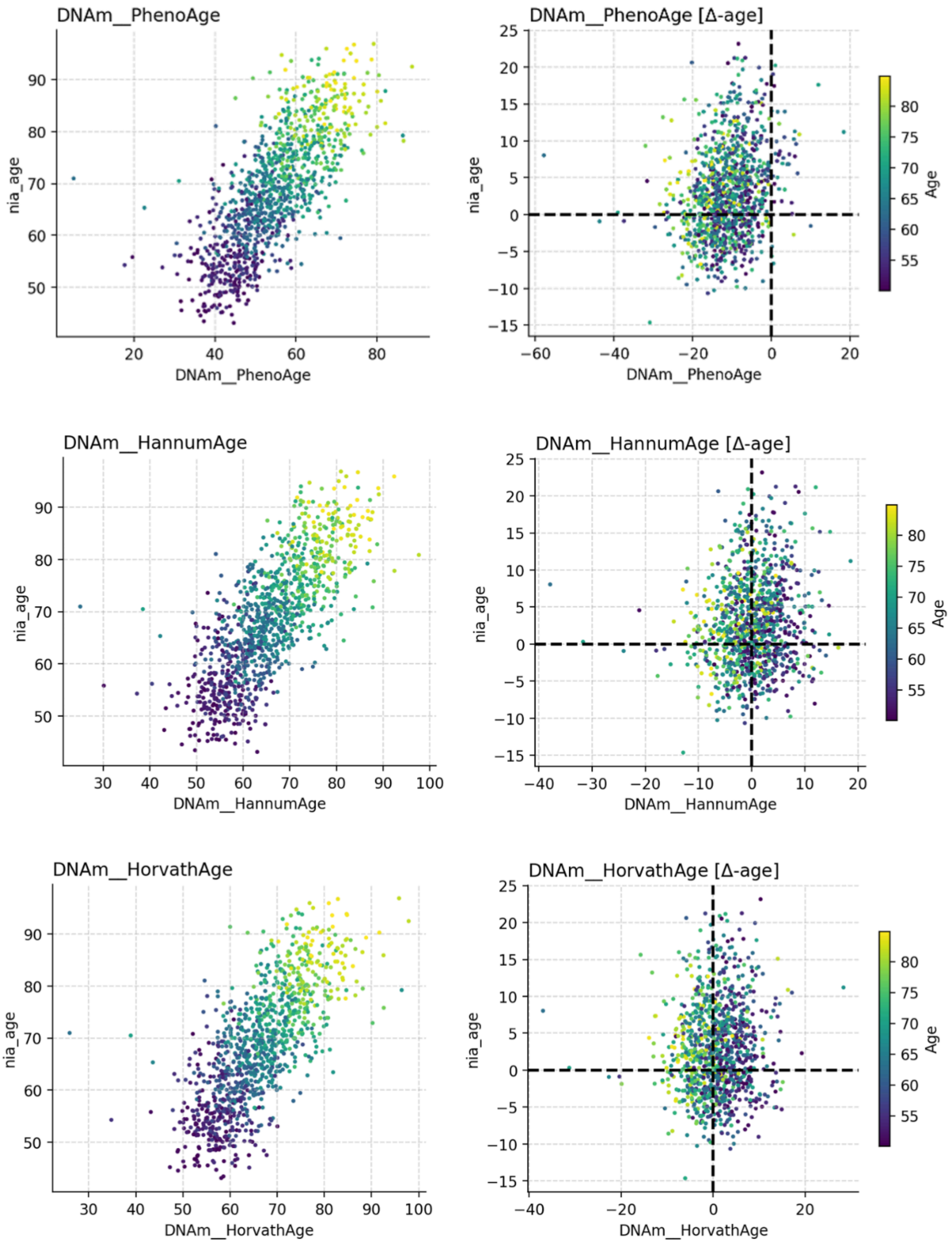
DNAm__PhenoAge
nia_age
DNAm__PhenoAge
DNAm__PhenoAge [Δ-age]
nia_age
DNAm__PhenoAge
Age
DNAm__HannumAge
nia_age
DNAm__HannumAge
DNAm__HannumAge [Δ-age]
nia_age
DNAm__HannumAge
Age
DNAm__HorvathAge
nia_age
DNAm__HorvathAge
DNAm__HorvathAge [Δ-age]
nia_age
DNAm__HorvathAge
Age

**Figure 2.** Delta-age deviations are less correlated with chronological age in NiaAge compared to existing age-clocks, a desirable property

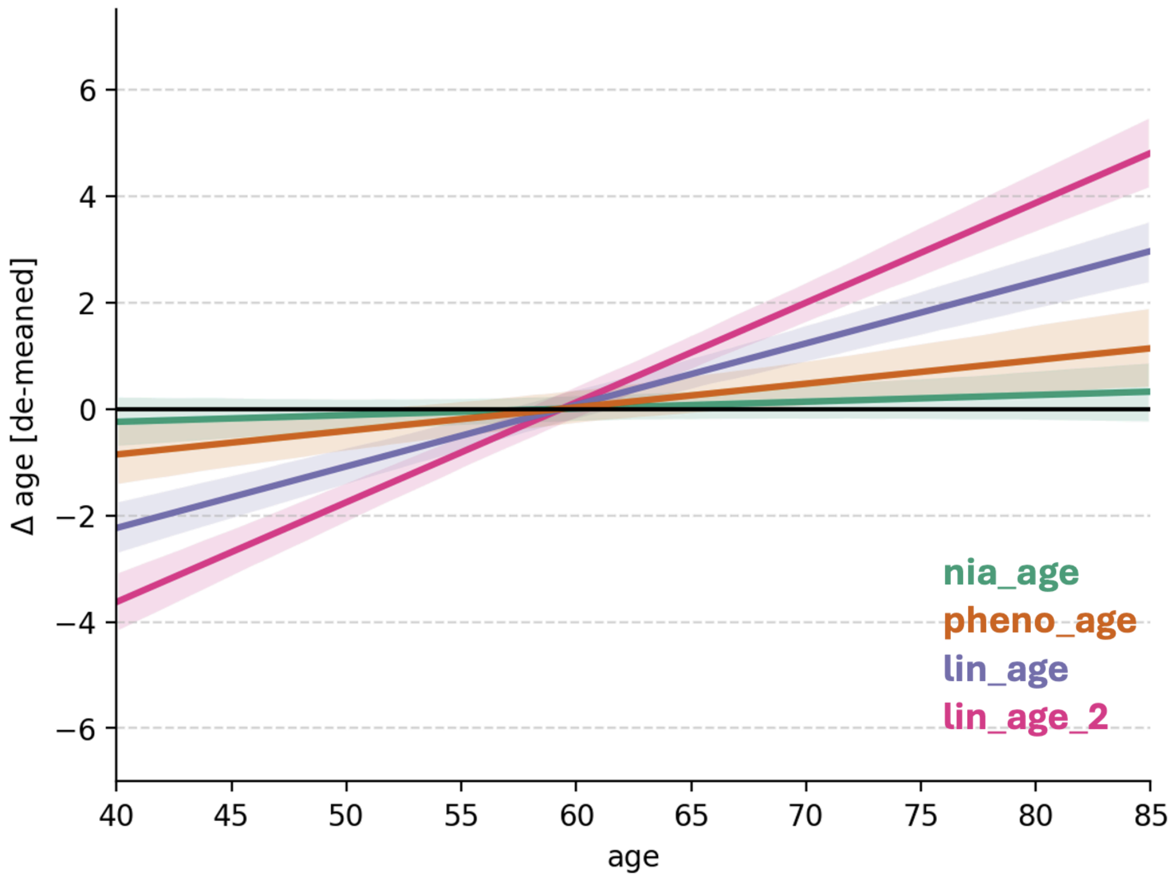


Regression lines (± standard error) predicting delta-age from chronological age. In existing age-clocks, older adults tend to exhibit higher delta-age deviations on average compared to younger adults. By explicitly re-referencing NiaAge biomarker contributions to age-expected values, NiaAge exhibits the desired property whereby delta-age deviations are not dependent on chronological age. NiaAge: $r = .02$, $p = .19$; LinAge: $r = .22$, $p <$ 1e-25; LinAge2: $r = .32$, $p <$ 1e-25; PhenoAge: $r = .07$, $p =$1e-4.

**Figure 3.** Mortality prediction in training and testing data-sets across age-clocks.

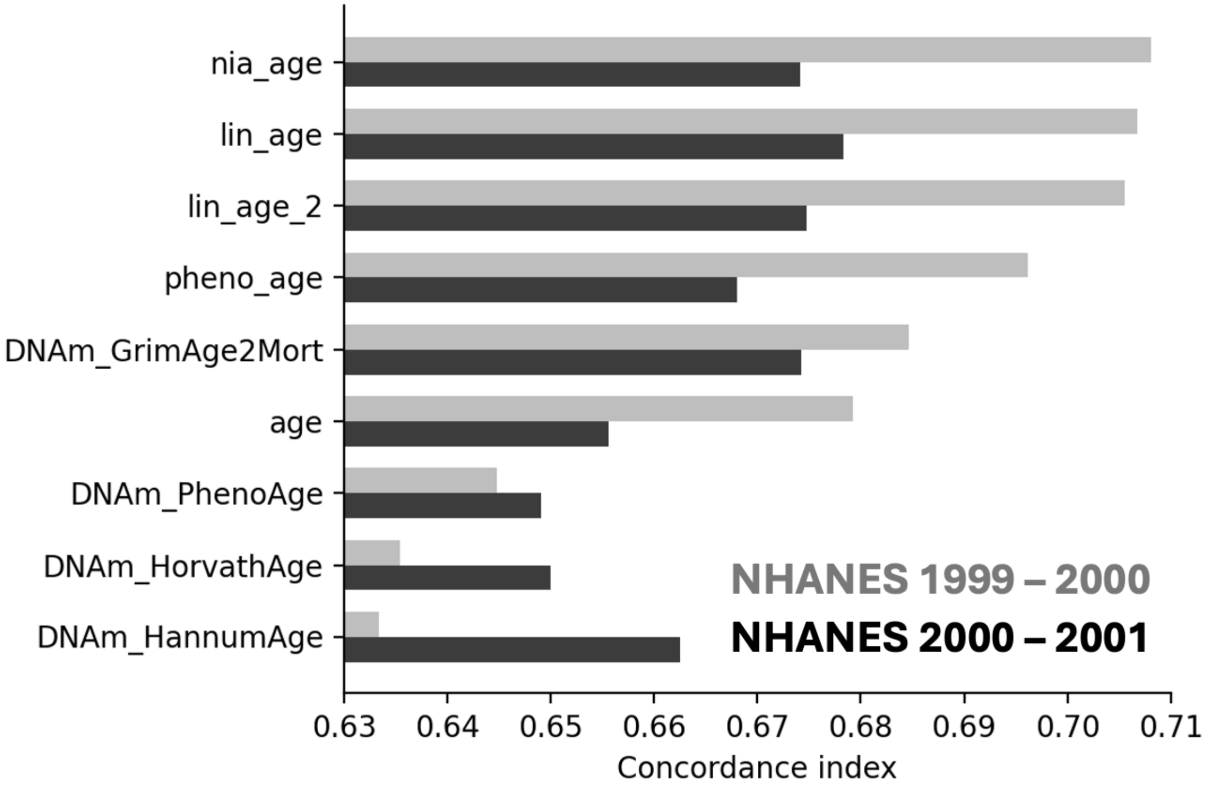


Barplot showing the concordance index (a measure of prediction accuracy) for each age-clock, in both training and testing NHANES data-sets.
